# Bibliometric Analysis of COVID-19 in the Context of Migration Health: A Study Protocol

**DOI:** 10.1101/2020.07.09.20149401

**Authors:** Sweetmavourneen Pernitez-Agan, Mary Ann Bautista, Janice Lopez, Margaret Sampson, Kolitha Wickramage

## Abstract

**Introduction:** Human mobility has been pivotal to the spread of COVID-19 through travel and migration. To mitigate the spread, most countries have imposed strict travel restrictions that have severely affected both the wellbeing and livelihoods of many migrant and mobile populations (both internally and internationally), particularly those from impoverished communities, those affected by humanitarian crises, including populations displaced and/or living in camps and camp-like settings. The need to include migrants (both regular and irregular or ‘undocumented’) in national strategic response plans for disease prevention and control has been increasingly recognized. Better understanding of the existing scientific evidence in migration health is crucial in designing effective response measures. In this paper, we present a protocol for a bibliometric analysis of scientific publications on COVID-19 and migration health. Expected study findings aim to provide valuable information to support evidence mapping on COVID-19 and migration health, particularly the identification of important research gaps.

**Methods and analysis:** Using Elsevier’s Scopus abstract and citation database, a comprehensive search strategy will be applied to map scientific publications on COVID-19 and migration health. The current analysis will focus on research published from 1 January 2020 to 4 May 2020. The search query on migration health will largely focus on migration, migrant and human mobility-related terms. Three reviewers will screen publications for eligibility. The extracted bibliographic information will be analysed to determine the dominant research themes, country coverage and migrant groups. Collaboration networks will be analysed using VosViewer, a network analysis software. A deep dive on dominant research themes or migrant health-related topics will be done by creating visualization network maps of keywords from the retrieved publications.

**Ethics and dissemination:** This analysis will draw on publicly available data and does not directly involve human participants; ethics review is not required.

## INTRODUCTION

### Migration health and COVID-19

Human mobility has played a central role in the spread of the coronavirus disease 2019 (COVID-19) [1, 2]. Movement of people from countries with known COVID-19 cases has emerged as a key driver in the rapid spread of the disease worldwide [3-7]. Several governments issued travel restrictions or outright bans on the entry of persons from countries or areas with known cases of COVID-19 and/ or suspension of domestic and international flights, which limited internal and global movement [1, 8].

Migration, among other factors, is considered a social determinant of health [9]. The relationship between migration and health is complex, and its impact varies considerably across migrant groups, and from person to person within such groups. The process of migration exposes migrant groups to various “health risks through unsafe travel, exposure to diseases, limited access to health services, poor nutrition, psychosocial stressors, and harsh living and working conditions” [10, 11].

Mitigating a global pandemic requires equal access to health services, regardless of migration status. Although various measures have been implemented in the COVID-19 pandemic response, these measures largely focus on protecting the local communities in host countries [12, 13]. Addressing the health of migrants and mobile populations is important in ensuring global health security; “migration health is a shared responsibility with public health impacts that extend beyond national boundaries” [10].

### Impact of COVID-19 among migrant populations

The COVID-19 pandemic has affected migrant and mobile populations perhaps to a greater extent than the general population [14]. Factors that contribute to this vulnerability include: (1) Precarious working environment and poor living conditions (e.g. temporary shelters, refugee camps), particularly for low-waged migrant workers, refugees, asylum seekers, and internally displaced populations (IDPs); (2) Limited or no access to health care services due to the legal and practical barriers to healthcare; in general, migrants have also been excluded in national pandemic plans [15]; (3) Travel restrictions that lead to mass movement of migrant worker populations back to their homes (international and internal); (4) Economic impact of the pandemic to migrant workers; and (5) Increasing xenophobia against migrant populations due to importation risks [16-20]. Migrants and mobile populations are frequently neglected, stigmatized, and potentially face difficulties in accessing health services that are otherwise available to the general population. In the context of the Interim Guidance on scaling-up COVID-19 outbreak in readiness and response operations for camps and camp-like settings (developed by the International Federation of Red Cross and Red Crescent Societies (IFRC), IOM, United National High Commissioner for Refugees (UNHCR), and World Health Organization (WHO), people in humanitarian situations may include IDPs, host communities, asylum seekers, refugees and returnees, and migrants in similar situations [21].

### Global response strategies to COVID-19

IOM, as part of the Inter-Agency Standing Committee (IASC), and in partnership with WHO, other United Nations (UN) organizations and coordination groups as well as non-UN stakeholders, is assisting Member States (MS) and partners to prepare for and respond to COVID-19, with operational, technical and policy support. One of the priorities outlined in IOM’s COVID-19 Global Strategic Preparedness and Response Plan (SRP) involves supporting efforts that properly consider the cross-cutting humanitarian and development needs of migrants, IDPs, and other vulnerable populations in reducing COVID-19-related illness and deaths [22].

### Research mapping: A migration health research priority

Bibliometric analysis is the quantitative analysis of publications (e.g. research articles and books) using bibliographic data (i.e. author information, citation, and publication information) to produce measures of ‘research productivity’ (i.e. number of publications), ‘research impact’ (i.e. citation counts, journal impact factor, etc.), and national or international networks/ collaborations of authors/ researchers, institutions/ organizations, and country/author affiliation). Although the bibliometric method relies on meta-data, it does not involve analysis and interpretation of the content of individual research publications. It has been firmly established as a scientific specialty and an integral part of research evaluation methodology. It provides useful information on the growth, impact, gaps, and trends of research publications within a particular field or discipline [23-25].

The 2^nd^ Global Consultation on Migrant Health (2017) recognized the need to “take stock of current research, map the existing landscape of published literature, identify areas of focus and gaps to better organize a global research agenda on migration health” [26]. In this paper, we present the protocol for a bibliometric study that aims to identify and analyze research publications on COVID-19 focusing on migration, migrants and human mobility; specifically mapping research productivity on COVID-19 in the context of migration health by author, country, institution/ organization, health theme, and migrant topic (i.e. migrant type and country coverage). The findings from this study will provide useful information in enhancing the strategic response to COVID-19 and will contribute to improving efforts in the successful integration of different migrant groups into the national COVID-19 preparedness and response plans.

## METHODS

### Citation database

Scopus, a citation and abstract database of peer-reviewed literature developed by Elsevier, will be used to retrieve publications on COVID-19 and migration health. Scopus provides a comprehensive overview of global research output in different disciplines and covers 100 per cent of MEDLINE publications. The advantage of Scopus over other citation databases was extensively discussed in previous studies [23, 27, 28].

### Search strategy and selection criteria

#### Inclusion steps

Two search queries will be developed for COVID-19 and migration health. Identification and selection of keywords will be based on the review of the WHO COVID-19 repository [29] and other bibliometric studies on COVID-19 [31] and migration health [23].

Search terms and Boolean separators for COVID-19 include “covid*” OR “covid-19” OR “covid19” OR “SARS-CoV-2” OR “ncov” OR “2019-nCov” OR “2019nCov” OR “corona virus” OR “coronavirus” and a combination of COVID-19 specific terms.

The search queries for COVID-19 and migration (health) will be combined using the Boolean operator “AND.” The search results of these queries will contain all publications on COVID-19 with ‘migration’, ‘migrant’ and ‘human mobility’ terms. It should be noted that while the analysis intends to capture publications on COVID-19 and “migration health”, the search query on migration health will largely focus on migration-, migrant- and human mobility-related terms as the health aspects of migration are effectively subsumed in the COVID-19 search query.

#### Exclusion steps

The following steps will be applied to the search strategy to eliminate irrelevant publications or false positive results.

- Restrict the publication year to 2020.
- Exclude publications indexed in irrelevant subject areas (e.g. Veterinary) after careful review of the retrieved publications. (NB: Scopus classifies retrieved publications based on the field and scope of the sources or publishing journal).
- Exclude publications with irrelevant or out of scope topics.
  a. The assigned reviewers will screen the title and abstract (if available) screening of the retrieved publications using MS Excel.
  b. The assigned reviewers will discuss among themselves the publications identified as “uncertain” and “excluded” until a consensus is reached on whether to include or exclude a publication.
- Exclude confirmed duplicates; duplicates will be identified using MS Excel and EndNote based on the following parameters: author names; publication title; source title; and, volume and issue number.

#### Validity of the search strategy

In each step of the search query, the search results will be reviewed to check the publication yield. The search strategy will be adjusted if known relevant publications are not captured in the search. Careful screening of the title and abstract will be done to ensure the validity of search results. The methodological rigor of the study will be reviewed and validated by a bibliometric analysis expert.

#### Data items and data extraction

The Scopus search output will be exported into several formats including CSV (for screening, classification, analysis, and visualization), RIS (for screening duplicates in EndNote), and BibTex (for analysis). All fields will be exported including the broad categories citation information, bibliographic information, abstract and keywords, funding details (where available) and cited references.

Bibliometric information that will be recorded from the online Scopus analysis include the following:

- Author names (with number of publications by author)
- Source title (with number of citations by source)
- Institution or organization name (with number of publications by institution/ organization)
- Country name from author affiliation address in Scopus (with number of publications by country)
- Publication type (with number of publications by type)
- Subject area (as defined by Scopus)
- Author and index keywords
- Funding source (number of publications by source)

#### Bibliometric analysis

Scopus and Biblioshiny will be used to analyze bibliometric information including authors, citations, publications, and sources (or journals). Biblioshiny is an open source web-interfaced bibliometrics tool that uses the R programme, a statistical software package [32]. Scopus has a built-in analysis function that can generate a list of leading publications, sources (or journals), authors, country author affiliations, institutions or organizations, and aggregates of publication types, and subject areas. Further analysis can be done on the leading authors, publications, and sources (or journals) using the profile feature available in Scopus. These two bibliometric tools provide the number of citations received for each publication and allow sorting of publications based on the number of citations. [33]

#### Network visualization mapping

VOSviewer version 1.6.15 [34], a software tool for constructing and visualizing bibliometric networks, will be utilized to analyze and visualize the networks of co-authorship relations among authors, countries, and institutions, and co-occurrence relations between keywords. To present a clean map, VOSviewer thesaurus files will be prepared to standardize terms and exclude generic and out-of-scope terms.

All network maps will be created using the network visualization option in the VOSviewer main panel. Table 1 lists the bibliographic data that will be used for the four visualization network maps (i.e. authorship, institution, country, and keyword networks) [35]. Items in networks are represented by circles. A network consists of a set of items linked together by lines. A network map contains at least one cluster; each cluster represented by a different color. The maps generated only present those networks with the largest set of links or connections. For example, the size of the circle represents the more frequently occurring keyword or the highest number of co-authored publications in the retrieved publication set from Scopus. The strength of links indicates the number of publications that two authors have co-authored (in the case of co-authorship links) or the number of publications in which two keywords occur together (in the case of co-occurrence links). The distance between two items in the visualization indicates the relatedness (i.e. co-authorship or co-occurrences) of items. The shorter the distance between two items, the stronger the relatedness. Colors represent clusters of items that are relatively strongly related to each other.

**Table 1.** Bibliographic data used in creating the visualization network map using VOSviewer

| Links* | Items** | Network |
| --- | --- | --- |
| Co-authorship | Institutions/ Organizations | Institution collaboration |
|  | Authors | Author collaboration |
|  | Countries | Country collaboration |
| Co-occurrences | Keywords | Keyword co-occurrences network |

\* A connection or a relation between two items in a visualization network map, represented by lines. \*\* Refers to the selected bibliographic data that will be used in the network mapping.

#### Research themes and subthemes

The authors will classify the retrieved publications into themes and subthemes reflective of the relevant migration health-related topics and IOM’s COVID-19 Strategic Preparedness and Response Plan (SPRP) [22].

Below is a description of each of the themes and subthemes:

1. Public health intervention *Description:* Publications that cover topics on any combination of program elements or strategies related to addressing COVID-19-related health concerns in different populations. *Subthemes:* Government measures; travel-related measures (e.g. travel restrictions and point-of-entry health screening); disease surveillance; community screening; case identification and management; contact tracing and management; personal protective measures (e.g. face masks and hand washing); social distancing measures (e.g. city lockdown and quarantine); environmental measure (e.g. disinfection of public spaces, indoor air quality measures); health education; health promotion (e.g. awareness campaign); and, mental health support.
2. Health system capacity *Description:* Publications involving topics related to health system capacity. *Subthemes:* health systems; leadership and governance (e.g. legal frameworks); health workforce (i.e. adequacy and capacity); medical products, vaccine, and technology (i.e. availability and procurement); health facility information (e.g. patient database); health financing; service delivery; continuity of routine health programmes (e.g. maternal, child and reproductive health); and, coordination and partnerships (i.e. coordination among relevant actors to support the pandemic response).
3. Clinical management *Description:* Publications that cover topics on characterizing the disease based on observing actual patients, treatment algorithms, management of patients and preventing and controlling infections (i.e. patient level management) *Subthemes:* clinical examination; clinical characterization (i.e. symptoms, characteristics, and disease progression); clinical guidance; and, clinical management (e.g. patient level supportive treatment)
4. Candidate therapeutics and vaccine *Description:* Publications that cover topics on the use of potential therapeutics (existing therapeutics) and vaccines (development of new ones). *Subthemes:* Potential therapeutics; and, candidate vaccines
5. Disease epidemiology and mathematical modelling *Description:* Publications that cover topics on disease etiology, distribution, and potential determinants (may include epidemiological approaches or other mathematical modelling). *Subthemes:* disease etiology (e.g. virus origin, viral structure); disease transmission; disease distribution (e.g. frequency, pattern); disease determinants (e.g. exposure variables and importation risks); and, mathematical modelling
6. Diagnostic and testing strategies *Description:* Publications that cover topics on diagnostic procedures and tests for COVID-19. *Subthemes:* diagnostic procedures; and, COVID-19 tests
7. Impact assessment and policy analysis *Description:* Publications that cover topics on socio-economic and health impact of COVID-19, health policy analysis and health diplomacy. *Subthemes:* cost-effective analysis; socio-economic analysis; and, health impact analysis
8. Migrant specific themes *Description:* Publications that cover topics specific to migrant support services. *Subthemes:* camp coordination and management (i.e. refugee camp and displacement site level); and, migrant protection (i.e. support services or programmes for migrant health protection)

Search terms will be identified for each theme and subtheme based on careful review of related publications. The search terms will be applied in the publication title and validated by title and abstract (if needed) screening.

#### Migration and mobility topics and country coverage

Migrants and mobile populations (e.g. international students, tourists, migrant workers, immigrants, refugees, and travelers) will be identified in the set of retrieved publications. Publications with no specific migrant or mobile population group will be screened for terms relevant to ‘human mobility’ (i.e. travel, transportation, and any form of movement within and across countries). The search terms will be applied in the publication title and abstract (if needed and available).

Further, ‘country coverage’ will be determined by scanning the title and abstract (if needed and available) of the publications. The term ‘country coverage’ refers to the country or countries identified as the main topic of the publication (i.e. where the study was implemented, where the data used in the study was obtained, or the country of citizenship and/or origin of the study population).

#### Database limitations

The depth and breadth of the findings from bibliometric analysis will depend on the information available in Scopus and the search strategy applied. Noted limitations inherent in a bibliometric study are as follows: (1) Relevant publications might be missed, particularly, those published in preprint servers. Research papers in the online preprint servers are not indexed in Scopus as these have yet to be peer-reviewed or accepted by traditional academic journals. Nevertheless, articles-in-press (i.e. pre-published versions of accepted research articles) are included in Scopus. (2) New publications might be missed due to time lag in the Scopus indexing (NB: Published articles are estimated to appear in Scopus within three to four weeks of publication on the publisher’s website). However, publishers and database producers are handling COVID-19 articles on a priority basis. (3) Bibliometrics only measures impact in terms of research productivity and not the research quality. (4) Search results reflect how the publication information was recorded and presented in Scopus. For example, active institutions, author names, and countries with different spellings will be spread out in the results.

## Data Availability

Data pertaining to the study protocol could be retrieved using the proposed Scopus and search
strategy.

## AUTHOR’S CONTRIBUTION

KW conceived the idea for the study. KW, SA, MB and JL were involved in the development of the study protocol. SA prepared the first draft of the study protocol and all authors critically reviewed, revised and approved the subsequent and final version. MS reviewed and validated the methodology of the study protocol and provided feedback on the search strategy development.

## ACKNOWLEDGEMENT

None.

